# Temporal trends and spatial variation in nontuberculous mycobacterial incidence among First Nations people in Queensland, Australia

**DOI:** 10.64898/2026.07.12.26357887

**Authors:** Melinda M. Ashcroft, Felicia Goh, Atha Rifqia Pradana, Scott C. Bell, Rachel M. Thomson

## Abstract

**Background:** Nontuberculous mycobacteria (NTM) are environmental pathogens causing pulmonary and extrapulmonary infections. First Nations people in Australia experience higher burdens of communicable diseases, comorbidities, and systemic barriers to care, increasing NTM risk. This study examined the incidence and spatial distribution of NTM infections in First Nations people in Queensland.

**Methods:** A retrospective longitudinal analysis was conducted using NTM notifications from the Queensland Health Notifiable Conditions Database, stratified by Indigenous status. Incidence was calculated using population denominators and Indigenous Region boundaries, with direct rate comparisons between 2011 and 2024.

**Results:** Between 2001 and 2024, 717 NTM notifications were recorded from 606 First Nations people, with a significant male predominance among those aged 30–44 years (χ²=21.63, *P*<0.0001). NTM incidence was higher in 2024 than in 2011, increasing from 4.6 to 26.3 per 100,000 (incidence rate ratio (IRR): 5.74, 95% confidence interval (CI): 2.97–11.08, *P*<0.0001). Although incidence in 2024 was lower than in the non-Indigenous population (35.81 per 100 000), the rate of increase was 4.4 times greater. Pulmonary infections predominated (569/717, 79.36%) and were more frequent in 2024 than in 2011 (IRR: 6.95, 95% CI: 3.00–19.75, *P*<0.0001). Extrapulmonary incidence increased by 140% from 2020 primarily due to an outbreak of *Mycobacterium abscessus* infections among incarcerated First Nations males. Marked geospatial heterogeneity was observed, with the greatest increases in incidence in the Brisbane, Rockhampton, Townsville–Mackay, and Cairns–Atherton Indigenous Regions (*P*<0.001).

**Conclusions:** NTM incidence among First Nations people in Queensland has increased substantially, with a faster rate of rise than in the non-Indigenous population despite lower absolute incidence, consistent with under-ascertainment. These findings highlight gaps in detection and diagnostic access, alongside heterogeneous geographic and outbreak-associated transmission dynamics. Strengthening culturally appropriate surveillance and improving access to timely diagnosis are required to better define disease burden and inform targeted clinical and public health responses in First Nations and other underserved populations.

## INTRODUCTION

Nontuberculous mycobacteria (NTM) are environmental pathogens that are an emerging but under-recognised public health threat [1]. Infection may range from colonisation and transient infections to severe invasive pulmonary and extrapulmonary disease. Immunocompromised individuals or those with structural lung diseases are at increased risk for NTM pulmonary disease (NTM-PD) [2]. Extrapulmonary infections include disseminated disease and localised infections of the skin, soft tissue, and other organs [3].

NTM infections are often underdiagnosed and frequently confused with *Mycobacterium tuberculosis* Complex infections, yet globally, incidence is increasing [4]. In Australia, NTM infections exceed those caused by Tuberculosis (TB) [5]. In Queensland, Australia, NTM are a notifiable infection under the Queensland Public Health Act (2005) [6], and are recorded in the Queensland Health Notifiable Conditions (NoCS) database [7]. Queensland encompasses diverse climate zones and is sparsely populated, with remote inland communities and major urban centres concentrated along the east coast.

Queensland has the second largest population of Aboriginal and Torres Strait Islander peoples in Australia (273,119 people in 2021, 5.3% of Queensland’s resident population [8]), hereafter referred to as First Nations people. First Nations people live across urban, regional, and very remote settings throughout Queensland, resulting in heterogeneous access to health services and diagnostic pathways. They experience a disproportionate burden of communicable and respiratory diseases, shorter life expectancy, marked socioeconomic disadvantage, and systemic barriers to healthcare access [9]. They also have higher rates of smoking [10] and chronic respiratory diseases like bronchiectasis and Chronic Obstructive Pulmonary Disease (COPD) [11], which are known NTM risk factors [12]. These disparities are compounded by broader structural determinants, including limited access to health services, household overcrowding, adverse environmental exposures, and increased incarceration rates [9] (comprising 36% of the Australian adult prison population [13]). Similar disparities in infectious disease burden and access to care are observed among Indigenous and underserved populations globally [14, 15], highlighting the broader relevance of understanding NTM epidemiology in these settings.

Previous work has reported rising NTM incidence in the overall Queensland population [16–18] and a substantial burden of NTM disease in the Northern Territory [19, 20]. However, information on NTM among First Nations people in Australia remains scarce. Population-level epidemiological data on NTM among First Nations or Indigenous populations are extremely limited globally, with existing studies largely restricted to small regional or cohort-based analyses and lacking systematic surveillance-based data. This study aimed to quantify temporal trends and spatial variation in NTM incidence among First Nations people in Queensland, Australia.

## METHODS

### NTM Notifications

We obtained all NTM notifications from the Queensland Health NoCS database between 2001–2024. All public hospital mycobacterial cultures are sent to the Queensland Mycobacterial Reference Laboratory (QMRL) for speciation; private laboratories also reported positive cultures and either speciate internally or send isolates to QMRL for speciation [21]. Species identification methods changed over time. Matrix-Assisted Laser Desorption/Ionization Time-of-Flight (MALDI-TOF) was introduced in 2019, and in 2021 PCR genotyping databases were updated and *M. fortuitum spp.* was reported as *M. fortuitum* Group (MFG). For consistency, all *M. fortuitum*, *M. peregrinum*, and all *M. fortuitum* Complex notifications were renamed as *M. fortuitum* Group (MFG). From 2021, most notifications of *M. avium* Complex (MAC) were either reported only at the complex level or involved MAC species other than *M. avium* or *M. intracellulare*.

Notifications were classified by Indigenous status. Individuals identifying as Aboriginal, Torres Strait Islander, or both were classified as First Nations people; unknown status was assumed non-Indigenous. Erroneous notifications where the individual had an acid-fast bacillus-positive but NTM-negative sample were excluded (n=104). A new notification was recorded if a positive sample was received more than 12 months after the initial positive sample. Where multiple species were identified in a single individual, notifications were duplicated by species. Demographic data included age, sex, Indigenous status, sampling date, and residential address. Residential addresses were used to determine whether the individual was incarcerated at sampling date. Site of infection and specimen type were used to classify infection type.

### Population Data

Indigenous Regions (IREGs) are large Australian Bureau of Statistics (ABS)–defined areas used for Aboriginal and Torres Strait Islander populations reporting [22]. Eight IREGs cover Queensland. Due to changes in the reporting of First Nations people between the 2006 and 2011 Censuses, incidence calculations were restricted to 2011–2024. Incidence rates per 100,000 First Nations people were stratified by sex using ABS mid-year population estimates (2011–2024) [23]. Non-Indigenous estimates were calculated as the total Queensland population minus First Nations people. IREG-level population data was obtained from the 2011, 2016, and 2021 Censuses, and the IREG 2021 boundaries (Australian Statistical Geography Standard Edition 3) were downloaded as a shapefile from the ABS [24].

### Statistical analyses, geospatial distribution, and code

Statistical analyses and mapping were conducted in R v4.5.1 [25]. Summary statistics were generated, and group differences were assessed using chi-square tests, and Wilcoxon rank-sum tests, as appropriate. Incidence rate ratios (IRRs) comparing 2011 to 2024 were estimated using unconditional maximum likelihood estimation with Wald confidence intervals. Residential addresses (n=720) or suburbs and localities (SALs; n=51) were geocoded in June 2025 using the *arcgis* method within *tidygeocoder* v1.0.5 [26]; SALs were assigned centroid coordinates. IREG-specific IRRs were estimated using multivariable Poisson regression with population included as an offset and stratified by Indigenous Region and Census year (2011, 2016, and 2021). Geographic variation in NTM notifications was examined using two-dimensional kernel density estimation, and geographic variation in incidence was examined using choropleth mapping of IREG-specific IRRs by Census year.

## RESULTS

### NTM incidence significantly increased among First Nations people in Queensland

Between 2001-2024, a total of 25,198 NTM notifications were recorded in Queensland, of which 717 (2.85%) were from 606 First Nations people. Among these individuals, 11.6% (70/606) had multiple notifications (median=2, range=2–7). The median age at sample collection was 55 years (IQR: 38-66), with 54.1% of notifications in males (Table 1). Overall, the male predominance was primarily attributable to significantly higher incidence among males aged 30–44 years compared with other age groups (χ²=21.63, *P*<0.0001). Paediatric infections (≤16 years) accounted for 9.2% (66/717) of notifications, with a median age of 11 years (IQR: 4–14); 35 cases occurred in females and 31 males.

**Table 1.** Demographics of nontuberculous mycobacterial infection notifications in First Nations People in Queensland, 2001–2024^a^.

| Characteristic | Pulmonary | Extrapulmonary | Unspecified | TOTAL |
| --- | --- | --- | --- | --- |
| **Sex** |  |  |  |  |
| Female (F) | 265 | 59 | 5 | 329 |
| Male (M) | 304 | 73 | 11 | 388 |
| **Indigenous Status** |  |  |  |  |
| Aboriginal | 450 (213F, 237M) | 109 (49F, 60M) | 11 (4F, 7M) | 570 (266F, 304M) |
| Torres Strait Islander | 87 (43F, 44M) | 13 (5F, 8M) | 4 (1F, 3M) | 104 (49F, 55M) |
| Both Aboriginal and Torres Strait Islander | 32 (9F, 23M) | 10 (5F, 5M) | 1 (0F, 1M) | 43 (14F, 29M) |
| **Age** |  |  |  |  |
| 0-14 | 25 (11F, 14M) | 27 (17F, 10M) | 2 (1F, 1M) | 54 (29F, 25M) |
| 15-29 | 51 (27F, 24M) | 19 (7F, 12M) | 3 (0F, 3M) | 73 (34F, 39M) |
| 30-44 | 69 (21F, 48M) | 37 (7F, 30M) | 5 (3F, 2M) | 111 (31F, 80M) |
| 45-59 | 159 (82F, 77M) | 32 (17F, 15M) | 4 (1F, 3M) | 195 (100F, 95M) |
| 60-74 | 198 (95F, 103M) | 16 (10F, 6M) | 2 (0F, 2M) | 216 (105F, 111M) |
| 75+ | 67 (29F, 38M) | 1 (1F, 0M) | 0 | 68 (30F, 38M) |
| **IREG** |  |  |  |  |
| Brisbane | 175 (94F, 81M) | 53 (22F, 31M) | 4 (2F, 2M) | 232 (118F, 114M) |
| Cairns–Atherton | 78 (30F, 48M) | 21 (11F, 10M) | 1 (0F, 1M) | 100 (41F, 59M) |
| Cape York | 38 (10F, 28M) | 7 (4F, 3M) | 0 | 45 (14F, 31M) |
| Mount Isa | 36 (16F, 20M) | 4 (3F, 1M) | 0 | 40 (19F, 21M) |
| Rockhampton | 80 (42F, 38M) | 16 (6F, 10M) | 4 (0F, 4M) | 100 (48F, 52M) |
| Toowoomba–Roma | 52 (25F, 27M) | 7 (6F, 1M) | 0 | 59 (31F, 28M) |
| Torres Strait | 46 (18F, 28M) | 5 (2F, 3M) | 1 (1F, 0M) | 52 (21F, 31M) |
| Townsville–Mackay | 64 (30F, 24M) | 19 (5F, 14M) | 6 (2F, 4M) | 89 (27F, 52M) |
| **TOTAL** | **569** | **132** | **16** | **717** |

<sup>a</sup>For notifications where the individual had multiple positive cultures over time, only one isolate per year was included. Notifications where the individual had multiple species were duplicated once per species.

Incidence in the non-Indigenous population was significantly higher in 2024 than in 2011, increasing from 17.33 to 35.81 per 100,000 (IRR: 2.07, 95% Confidence Interval (CI): 1.90–2.25, *P*<0.0001). For First Nations people, notifications increased from 2 in 2001 to 77 in 2024. Further, incidence among First Nations people was also significantly higher in 2024 than in 2011, increasing from 4.6 to 26.3 per 100,000, corresponding to a 5.7-fold increase (IRR: 5.74, 95% CI: 2.97–11.08, *P*<0.0001; Figure 1). Notably, the relative rate of increase was 4.4 times greater in First Nations people.

**Figure 1.**
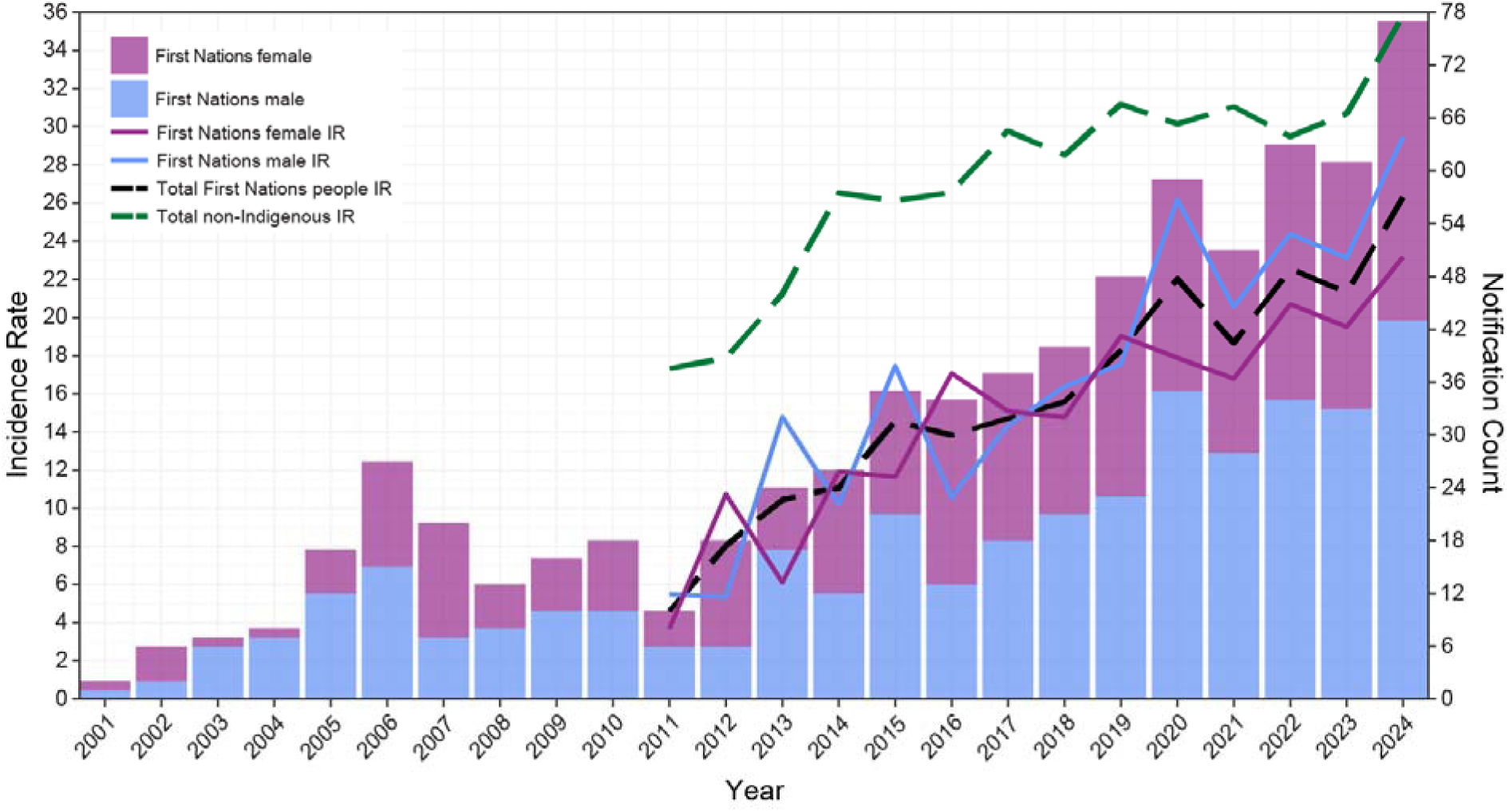
Nontuberculous mycobacterial (NTM) infection incidence (per 100,000 First Nations people) and NTM notification counts in First Nations people in Queensland, Australia. Incidence Rates (IR) are shown as line graphs, with the bar chart representing notification counts. Green dashed line shows the incidence rate per 100,000 non-Indigenous population.

### Pulmonary and extrapulmonary NTM incidence increased over time in First Nations people in Queensland

Infection type could be classified for 701 of the 717 NTM notifications (97.8%). Pulmonary infections were 4.3 times more common, with 569 notifications (79.4%), compared to 132 extrapulmonary infections (18.4%); 16 notifications (2.2%) were unclassified. This distribution differed in the paediatric cohort, where 37 notifications (56.1%) were pulmonary and 27 (40.9%) extrapulmonary (χ²=19.43, *P*<0.0001); two were unclassified. Among paediatric cases, median age varied significantly by infection type (W=863, *P*<0.0001), with pulmonary infections occurring at a median age of 13 years (IQR: 11–15; 20 males, 17 females) compared to 4 years for extrapulmonary infections (IQR: 2–9; 10 males, 17 females). Further, there were significant differences in the sex distribution of paediatric extrapulmonary infections compared to the adult cohort (χ²=4.58, *P*<0.032), with females comprising a higher proportion of paediatric cases (17/27; 63%), while males were more common among adult cases (63/105; 60%). In all notifications in First Nations people, pulmonary infections were predominantly diagnosed from sputum samples (520/569, 91.4%) or bronchoalveolar lavages (45/569, 7.9%). Extrapulmonary infections were most often skin, soft tissue, or wound infections (92/132, 69.7%), followed by bone and joint infections (17/132, 12.9%).

Pulmonary NTM incidence was significantly higher in 2024 than in 2011, increasing from 2.75 to 19.13 per 100,000 First Nations people (IRR: 6.95, 95% CI: 3.00–19.75, *P*<0.0001; Figure 2A). A total of 304 notifications occurred in males and 265 in females. Incidence was significantly higher in 2024 than in 2011 for both sexes, with a more pronounced increase in females, rising by 974% (IRR: 10.74, 95% CI: 2.72–92.88, *P*<0.0001), compared with a 406% rise in males (IRR: 5.06, 95% CI: 1.76–19.89, *P*=0.0004).

**Figure 2.**
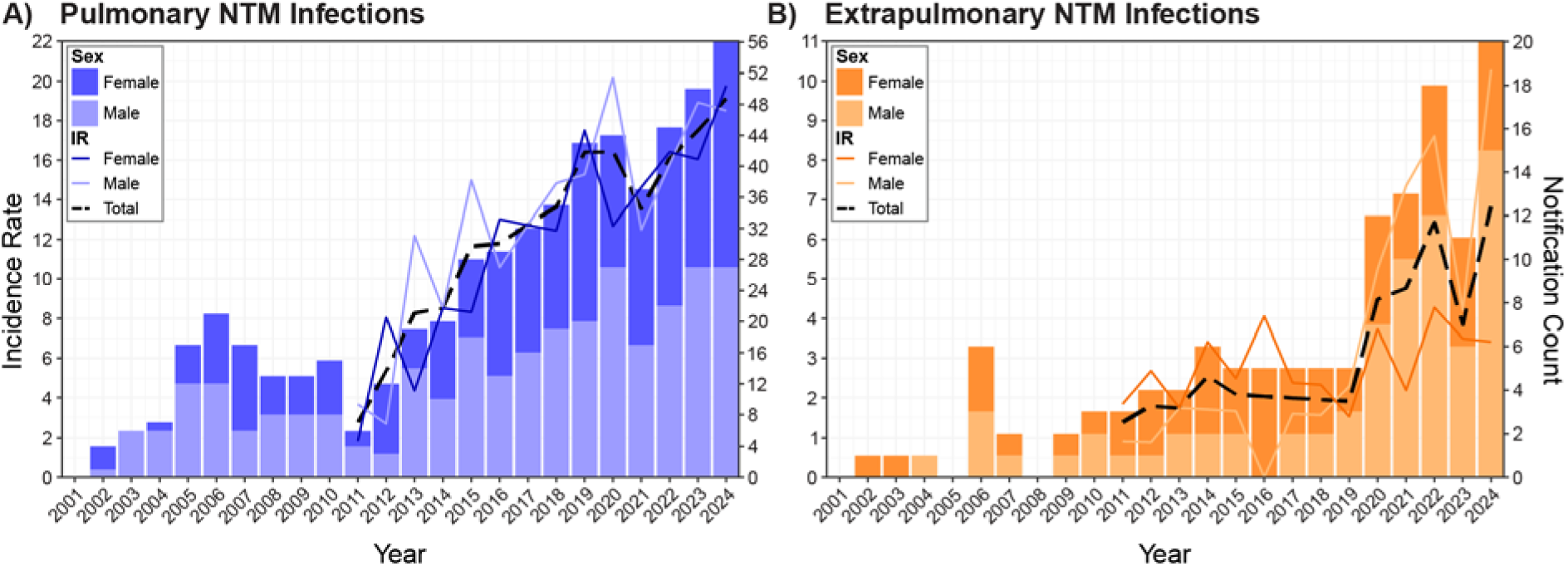
Nontuberculous mycobacterial (NTM) infection incidence (per 100,000 First Nations people) and notification counts in First Nations people in Queensland, Australia stratified by infection type and sex. A) Pulmonary NTM infections. B) Extrapulmonary NTM infections. Incidence rate (IR) is shown as a line graph, with the bar chart representing notification counts.

Extrapulmonary NTM incidence was also significantly higher in 2024 than in 2011, increasing from 1.38 to 6.83 per 100,000 First Nations people (IRR: 4.97, 95% CI: 1.47–26.1, *P*=0.003; Figure 2B). Incidence remained relatively stable between 2011 and 2019 (1.38–2.55 per 100,000) before a 140% increase in notifications in 2020 that persisted through 2024. This increase was predominantly in males, with incidence in males significantly higher in 2024 than in 2011, increasing by 1024% (IRR: 11.24, 95% CI: 1.73–473.04, *P*=0.002), compared with an 85% increase in females (IRR: 1.85, 95% CI: 0.30–19.44, *P*=0.49).

To explore the 2020 NTM outbreak in First Nations people in Queensland, we examined extrapulmonary NTM notifications from 2020-2024. Of the 74 notifications during this period, 26 (35.1%) notifications occurred from 26 incarcerated First Nations males. In contrast, in the same period among the non-Indigenous population, there were 875 extrapulmonary notifications, of which 41 (4.7%) were in incarcerated individuals (39 males, 2 females). Among the 26 notifications from incarcerated First Nations people (2020–2024), 20 were from skin, soft tissue, or wound infections (77%), three were from bone or joint infections (11.5%), and three were disseminated infections (11.5%). *M. abscessus* was identified in 92.3% (24/26) of these infections. Moreover, prior to 2020, there was only a single extrapulmonary NTM infection in incarcerated First Nations people in Queensland. When notifications from incarcerated individuals were excluded, extrapulmonary incidence dropped substantially. Crude incidence was higher in 2024 than in 2011 (3.42 vs. 1.38 per 100,000 First Nations people); however, this increase was not statistically significant (IRR: 0.40, 95% CI: 0.07–1.56, *P*=0.163; Supplementary Figure 1). Among extrapulmonary notifications from 2020 onward with incarcerated individuals excluded, repeat notifications were uncommon, and both the sex distribution and age profile were like the overall pattern (Table 1). These findings indicate that the observed increase was not driven by duplicate notifications or demographic shifts.

### NTM incidence in First Nations people varied across Queensland Indigenous Regions

Most NTM notifications were concentrated in densely populated areas, but marked geospatial heterogeneity was evident (Figure 3A). Beyond the major urban population centre of South-East Queensland, substantial notifications were recorded in the rural areas of Wide Bay and the Darling Downs, along the tropical east-coast, in Mount Isa, and in the Torres Strait. When adjusted for the underlying population of First Nations people, significant rate increases were identified in four of the eight Queensland Indigenous Regions, including Brisbane (IRR: 2.8, 95% CI: 2.31–3.37, *P* < 0.001), Rockhampton (IRR: 2.06, 95% CI: 1.67–2.52, *P*<0.001), Townsville–Mackay (IRR: 1.57, 95% CI: 1.25–1.96, *P*<0.001), and Cairns–Atherton (IRR: 1.45, 95% CI: 1.15–1.81, *P*=0.001; Figure 3B and Supplementary Table 1).

**Figure 3.**
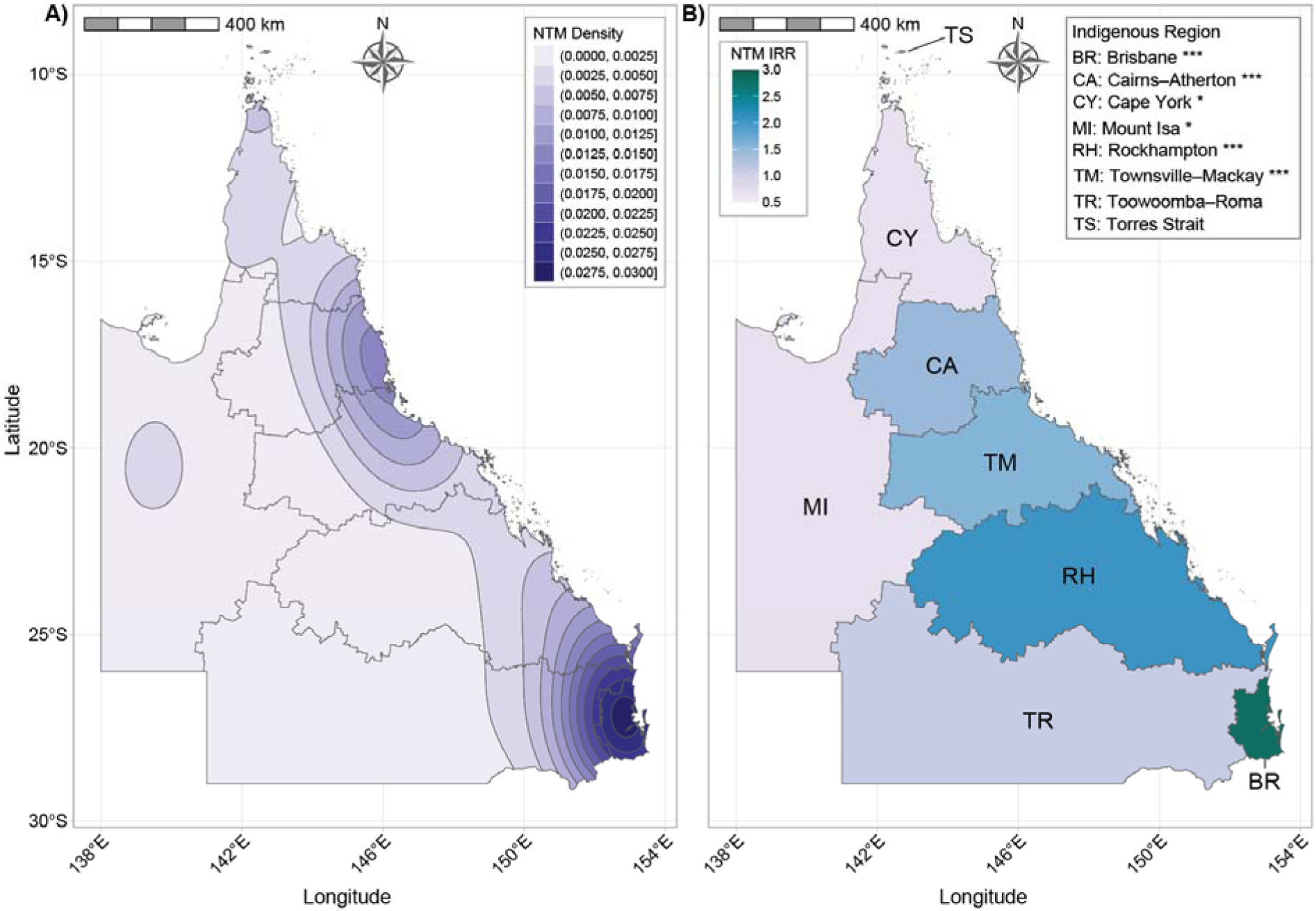
Geographic distribution and incidence rate ratios (IRRs) of nontuberculous mycobacterial (NTM) notifications in First Nations people in Queensland, Australia. A) Density of NTM notifications 2001–2024 with Queensland Indigenous Regions (IREGs) boundaries highlighted by grey borders. Light blue=lowest density, dark blue=highest density. B) Choropleth map of IRRs stratified by IREG and Census years 2011, 2016, and 2021. The darker the colour, the higher the rate of increase. Significance is indicated by asterisks where *=*P*≤0.05 and ***=*P*≤0.001.

### Five NTM species dominated across infection types

A total of 44 NTM species were responsible for NTM notifications in First Nations people in Queensland, with some recorded only at the complex level (e.g. *M. fortuitum* Group (MFG) and *M. avium* Complex (MAC)) or left unspeciated (not-typed; 86/717, 12%). Overall, five species or species groups accounted for 61.7% of all notifications: *M. intracellulare* (219/717, 30.5%), *M. abscessus* (93/717, 13%), MFG (73/717, 10.2%), *M. avium* (36/717, 5%), and *M. chelonae* (21/717, 2.9%) (Figure 4). Additionally, there were 30 MAC-level notifications, 28 slow grower-level notifications, 14 rapid grower-level notifications, and 30 other species had fewer than five notifications. The IRR increased for all top species except *M. chelonae*; however, incidence was highly variable over time (Supplementary Figure 2). Among the 70 First Nations people with multiple NTM notifications over the 24-year period, 25 had repeated isolation of the same species, 35 had two different species, and three had three different species, while species information was incomplete for seven individuals.

**Figure 4.**
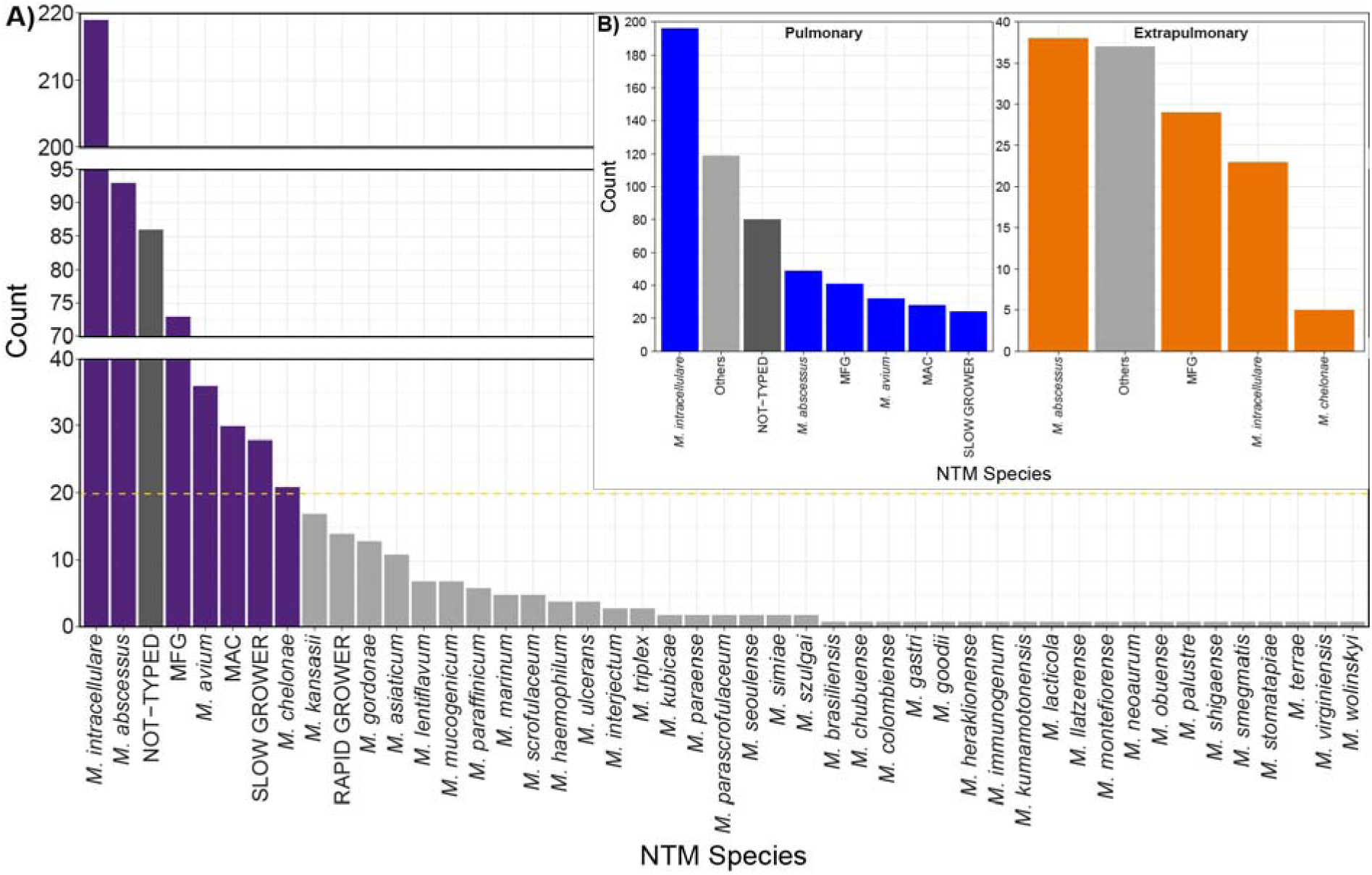
Spectrum of species isolated from NTM notifications in First Nations people in Queensland, Australia 2001-2024. A) All notifications. Species or groups that contained >20 isolates (indicated by the yellow dashed line) are coloured in dark purple. Rare and very rare species or groups (<20 isolates) are coloured in light grey. NTM isolates that were not-typed are shown in dark grey. Inset: B) Pulmonary isolates (left), Extrapulmonary isolates (right). Species or groups that had <20 notifications for pulmonary infections or <5 notifications for extrapulmonary infections were grouped as Others, shown in light grey. NTM isolates that were not-typed are shown in dark grey. Dominant species or groups were coloured blue for pulmonary infections and orange for extrapulmonary infections. MFG = *Mycobacterium fortuitum* Group, MAC = *Mycobacterium avium* Complex.

When infection type was considered, there were clear differences in NTM species patterns. In pulmonary infections, *M. intracellulare* dominated (196/569, 34.5%), followed by *M. abscessus* (n=49/569, 8.6%), MFG (n=41/569, 7.2%), and *M. avium* (n=32/569, 5.6%). In contrast, extrapulmonary infections were dominated by *M. abscessus* (38/132, 28.8%), MFG (29/132, 22%), and *M. intracellulare* (23/132, 17.4%). The high proportion of *M. abscessus* extrapulmonary notifications reflected the correctional centre outbreak. When notifications from incarcerated individuals (2020 onwards) were excluded, MFG became the dominant species, with *M. abscessus* dropping to third. Culture growth pattern also differed by infection type. Pulmonary infections were mostly caused by slow growers (364/569, 64%), whereas extrapulmonary infections were primarily due to rapid growers (82/132, 62.1%). In the non-Indigenous population, pulmonary infections were also dominated by the slow growers *M. intracellulare* (8274/20518, 40.2%) and *M. avium* (2138/20518, 10.4%). Extrapulmonary infections in the non-Indigenous population were similarly dominated by MFG (788/3353, 23.5%) and *M. abscessus* (566/3353, 16.9%). However, the rapid growers *M. marinum* (283/3353, 8.4%) and *M. chelonae* (282/3353, 8.4%) were more prominent than *M. intracellulare* (257/3353, 7.7%; Supplementary Table 2).

## DISCUSSION

This study demonstrates a 5.7-fold increase in NTM incidence among First Nations people in Queensland between 2011 and 2024, exceeding the incidence observed in the general Queensland population during earlier surveillance [18]. These findings align with global reports on rising NTM incidence associated with demographic shifts, increasing comorbidities rates, population ageing, environmental exposures, a changing climate, and improved recognition and diagnostics [4].

NTM incidence in 2024 was higher in the non-Indigenous population (35.8 vs. 26.3 per 100,000). However, the rate of increase was 4.4 times greater among First Nations people, indicating a rapidly growing burden despite lower absolute incidence, even after accounting for the extrapulmonary outbreak in incarcerated males. Similar patterns were reported in the Northern Territory, where First Nations people had lower rates of NTM-PD [20] and extrapulmonary infection [19] than the non-Indigenous population. These findings suggest that the lower observed incidence does not reflect a true lower burden, but rather under-ascertainment driven by systemic gaps in case detection. In Queensland, approximately 40% of First Nations people live in remote or very remote regions [27], where geographic isolation, transport constraints, limited availability of culturally appropriate healthcare, and diagnostic delays [9] hinder timely diagnosis. Detection is further complicated by the non-specific presentation of NTM-PD [28] and the normalisation of chronic cough as a routine rather than pathological symptom [29]. Communication barriers, discrimination, and mistrust arising from negative healthcare experiences may further deter presentation and follow-up [30]. Improved recognition of NTM in First Nations populations has important implications for clinical management, including earlier diagnosis, appropriate antimicrobial therapy, and differentiation from TB.

These systemic barriers obscure the true NTM burden while amplifying TB disparities. First Nations people in Australia experience substantially higher TB incidence than the non-Indigenous population [31, 32], a pattern also observed in New Zealand, Canada, and the United States [33]. This reflects, in part, enhanced case detection through Australia’s strategic plan for TB elimination [32] and increased exposure through cross-border travel from Papua New Guinea, where TB remains endemic [34]. Nonetheless, persistent social and health inequities continue to sustain elevated risk [35]. Access to diagnosis and healthcare engagement influence detection of both TB and NTM, and these processes are closely linked. Targeted TB surveillance increases case ascertainment, whereas the absence of equivalent pathways for NTM suppresses detection despite shared risk environments. Consequently, the contrast between lower recorded NTM and higher TB incidence reflects two manifestations of the same structural inequities [30], where detection is obscured in one case and clinical impact is magnified in the other.

Pulmonary NTM infections accounted for most notifications, consistent with previous Queensland studies [17]. Incidence increased sharply after 2011, unlike TB incidence, which remained relatively stable until the COVID-19 pandemic [36]. Prior TB infection and Bacille Calmette-Guérin (BCG) vaccination may confer partial cross-protection [37], although TB-related lung damage increases NTM-PD susceptibility [12]. Part of the increase in NTM incidence may reflect incidental detection during TB screening campaigns, particularly during the outbreaks in 2017-2020 and from 2023 [36]. In TB-focused settings, limited clinician awareness of NTM may delay diagnosis and follow-up, with surveillance data indicating frequent incidental NTM detections [38]. The temporary decline in pulmonary NTM in 2021 likely reflects behavioural changes during COVID-19 public health measures, which reduced opportunities for acquiring infectious diseases and decreased healthcare-seeking for non-urgent symptoms [39], consistent with parallel declines in TB notifications [40]. Concurrent service disruptions likely prolonged diagnostic delays, while fewer respiratory exacerbations [41] and reduced circulation of respiratory viruses [42] likely resulted in fewer sputum samples being collected and tested for NTM.

Extrapulmonary NTM infections remained uncommon until 2020, when an *M. abscessus* outbreak among incarcerated First Nations males led to a sharp increase. Incidence during this period was similar among incarcerated First Nations and non-Indigenous people (612.77 vs. 617.84 per 100,000 population), but the absolute impact was greater because First Nations people are imprisoned at substantially higher rates (2,435 vs. 174 per 100,000) [13]. After excluding incarcerated individuals, a modest increase in extrapulmonary incidence persisted, not explained by repeat notifications or demographic shifts. This suggests that factors beyond custodial exposure contribute to ongoing risk. High-risk injecting practices in prisons, particularly in the absence of needle-syringe programs, may also contribute to transmission [43]. These findings underscore the need for targeted harm-reduction and infection-control strategies in correctional facilities.

Age and sex also influence NTM incidence. In contrast to Queensland-wide data showing a female predominance in older adults [44], this study found a male predominance, particularly among those aged 30–44 years. This likely reflects greater occupational environmental exposure, higher smoking prevalence among First Nations males [45], and elevated rates of chronic respiratory diseases including bronchiectasis and COPD [46]. However, sex-related differences varied by infection type. Among extrapulmonary notifications, females comprised a higher proportion of paediatric cases (63%), where males predominated among adults (60%). In children, this aligns with previous reports and may reflect incomplete immune maturation and age-related behavioural exposures [47]. The lower proportion of extrapulmonary infections in First Nations children compared with non-Indigenous children may also relate to ongoing BCG vaccination and associated cross-protection [47]. In adults, extrapulmonary notifications from 2020 were inflated by the outbreak in incarcerated males. These patterns may also reflect underlying geographic and environmental factors that shape NTM distribution.

Marked geospatial heterogeneity was observed, with clusters in South-East Queensland, the agricultural regions of Wide Bay and the Darling Downs, the tropical north-east coast, the Torres Strait, and the inland Mount Isa mining region. The largest increases occurred in the Brisbane, Rockhampton, Townsville–Mackay, and Cairns–Atherton Indigenous Regions, consistent with previous studies showing elevated incidence in South-East Queensland and the northern tropical coast [17, 18]. The persistence of clusters in agricultural and mining areas suggests environmental and occupational exposures. Higher incidence in warm, wet climates may reflect heavy rainfall and flooding, which increase exposure to NTM-containing aerosols [48, 49]. Frequent cyclones [50] likely amplify these conditions, particularly in coastal regions that already show high incidence.

NTM notifications were dominated by *M. intracellulare*, *M. abscessus*, MFG species, *M. avium*, and *M. chelonae*, consistent with previous Queensland studies [17, 18]. Similar findings in the Northern Territory [19] support regional concordance, and the predominance of MAC in pulmonary infections and RGM in extrapulmonary infections reflects global trends [51]. The increase in *M. abscessus* from 2020 was driven by the correctional-facilities outbreak. Recent work demonstrating *M. abscessus* in Queensland municipal drinking water distribution systems supports treated water as an environmental reservoir [52]. *M. intracellulare* was the most common species but showed high year-to-year variability, while *M. avium* had only a modest increase. In contrast MFG and *M. chelonae* showed low incidence without clear temporal trends. Although heterogeneity was observed across Queensland Indigenous Regions, small sample sizes limited identification of species-specific hotspots. Species distributions in the non-Indigenous population however showed a different hierarchy, particularly for extrapulmonary infections where rapid growers like MFG, *M. abscessus*, *M. marinum*, and *M. chelonae* comprised a greater proportion. These contrasts may reflect variations in environmental contact, socioeconomic and occupational differences, and more consistent diagnostic follow-up in non-Indigenous patients.

This study has several limitations. NTM notification data capture positive cultures but cannot distinguish between contamination, colonisation, transient infection, or clinically significant disease. Our approach prioritised sensitivity over specificity, resulting in broader incidence estimates; for example, individuals with multiple positive cultures over time were recorded as one notification per species per year. Indigenous status may be misclassified or under-reported [53], leading to underestimation of incidence. Population denominators were derived from census data, limiting analyses to 2011 onward due to changes in ABS reporting methods [54]. Analyses were aggregated by ABS-defined Indigenous Regions to protect privacy, which, combined with small sample sizes, limited spatial resolution. Lastly, many species had fewer than five notifications, most appearing after 2019, which may partly reflect changes in QMRL laboratory methods, including the introduction of MALDI-TOF speciation in 2019 and genotyping database updates in 2021 that enabled identification of rarer species.

## CONCLUSION

This population-level study shows a 5.7-fold rise in NTM incidence among First Nations people in Queensland since 2011. Although incidence remains lower than in the non-Indigenous population, the more rapid increase indicates a growing burden likely compounded by under-ascertainment and environmental exposures. Pulmonary infections predominated, which may partly reflect increased detection through TB screening. The recent *M. abscessus* outbreak in correctional facilities and the higher proportion of extrapulmonary infections in First Nations children compared with adults further illustrate the diversity of clinical presentations and populations affected. These findings underscore the need for culturally safe surveillance, improved diagnostic access, and targeted interventions to reduce the growing burden of NTM infections in First Nations people.

## Data Availability

The data used in this study comprise statutory NTM notification records held by Queensland Health. These data cannot be shared publicly, including in de-identified form, because the rarity of NTM infections and the inclusion of First Nations status create a risk of re-identification. Data access is governed by Queensland, Australia legislation and specific public health and human research ethics approvals. Requests for access to aggregated data may be considered by Queensland Health, subject to ethical approval and data governance requirements.

## Abbreviations

ABS: Australian Bureau of Statistics
BCG: Bacille Calmette-Guérin
CI: confidence interval
COPD: Chronic Obstructive Pulmonary Disease
IQR: interquartile range
IREG: Indigenous Region
IRR: incidence rate ratio
MAC: *Mycobacterium avium* Complex
MALDI-TOF: Matrix-Assisted Laser Desorption/Ionization Time-of-Flight
MFG: *Mycobacterium fortuitum* Group
NoCS: Notifiable Conditions
NTM: Nontuberculous mycobacteria
PD: pulmonary disease
QMRL: Queensland Mycobacterial Reference Laboratory
SAL: suburbs and localities
TB: tuberculosis

## Acknowledgements

We would like to thank Bridget O’Connor from Queensland Health for her assistance in supplying the Notifiable Conditions Database.

## Author contributions

Conceptualisation: RMT, MMA, SCB. Funding acquisition: RMT. Supervision: RMT. Project administration: MMA, FG, RMT. Data curation: MMA, ARP, RMT. Methodology: MMA. Formal analysis: MMA, RMT. Writing – original draft: MMA, RMT. All authors reviewed the manuscript and approved the final version for submission.

## Availability of data and materials

The data used in this study comprise statutory NTM notification records held by Queensland Health. These data cannot be shared publicly, including in de-identified form, because the rarity of NTM infections and the inclusion of First Nations status create a risk of re-identification. Data access is governed by Queensland legislation and specific public health and human research ethics approvals. Requests for access to aggregated data may be considered by Queensland Health, subject to ethical approval and data governance requirements.

## Declarations

## Ethics approval

This study was approved by The Prince Charles Hospital Human Research Ethics Committee (HREC/15/QPCH/65). Approval to access NTM notification data was granted by Queensland Health (27466) under the Public Health Act 2005.

