## Supplementary Materials for "Temporal trends and spatial variation in nontuberculous mycobacterial incidence among First Nations people in Queensland, Australia"

**Supplementary Tables**

**Supplementary Table 1. Incidence rate ratios of nontuberculous mycobacterial infection notifications by Indigenous Region in First Nations people in Queensland.**

| **Characteristic** | **IRR** | **95% CI** | ***P*-Value** |
| --- | --- | --- | --- |
| Years^#^ | 1.15 | 1.13–1.17 | <0.001 |
| **IREG** | | | |
| Brisbane | 2.8 | 2.31–3.37 | <0.001 |
| Cairns–Atherton | 1.45 | 1.15–1.81 | 0.001 |
| Cape York | 0.73 | 0.53–0.97 | 0.038 |
| Mount Isa | 0.73 | 0.53–0.97 | 0.038 |
| Rockhampton | 2.06 | 1.67–2.52 | <0.001 |
| Toowoomba–Roma | 1.09 | 0.84–1.40 | 0.5 |
| Torres Strait | 1.09 | 0.83–1.41 | 0.5 |
| Townsville–Mackay | 1.57 | 1.25–1.96 | <0.001 |

^#^Incidence rate ratios (IRRs) were stratified by census years 2011, 2016, and 2021. IREG: Indigenous Region; CI: Confidence interval.

**Supplementary Table 2. Most frequently notified nontuberculous mycobacterial species among the non-Indigenous Queensland population, 2001**–**2024*.**

| **Species** | **Count (%)** | **Growth Rate** |
| --- | --- | --- |
| **Pulmonary** | | |
| *M. intracellulare* | 8274 (40.33) | Slow Grower |
| *M. avium* | 2138 (10.42) | Slow Grower |
| *M. abscessus* | 1740 (8.48) | Rapid Grower |
| MFG | 940 (3.70) | Rapid Grower |
| MAC | 562 (2.74) | Slow Grower |
| *M. gordonae* | 520 (2.43) | Slow Grower |
| *M. chelonae* | 374 (1.82) | Rapid Grower |
| **Extrapulmonary** | | |
| MFG | 787 (20.37) | Rapid Grower |
| *M. abscessus* | 566 (16.88) | Rapid Grower |
| *M. marinum* | 283 (8.44) | Slow Grower |
| *M. chelonae* | 282 (8.41) | Rapid Grower |
| *M. intracellulare* | 257 (7.66) | Slow Grower |
| *M. avium* | 144 (4.29) | Slow Grower |

*For notifications where the individual had multiple positive cultures over time, only one isolate per year was included. Notifications where the individual had multi-species infections were duplicated once per species. MFG = *Mycobacterium fortuitum* Group, MAC = *Mycobacterium avium* Complex.

**Supplementary Figures**

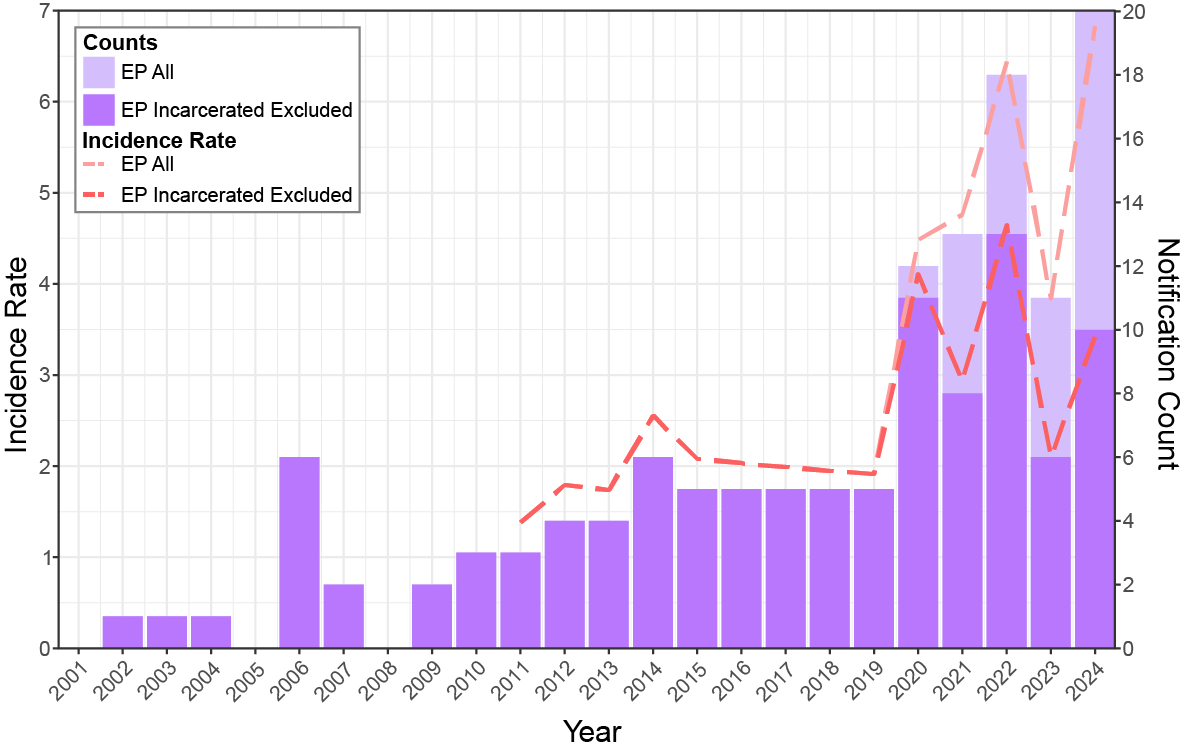

**Supplementary Figure 1. Extrapulmonary nontuberculous mycobacteria (NTM) incidence (per 100,000 First Nations people) and notification counts in First Nations people in Queensland.** EP All: All extrapulmonary NTM notifications in First Nations people. EP Incarcerated Excluded: extrapulmonary NTM notifications from First Nations people who listed a Queensland Correctional Centre as their place of residence were excluded.

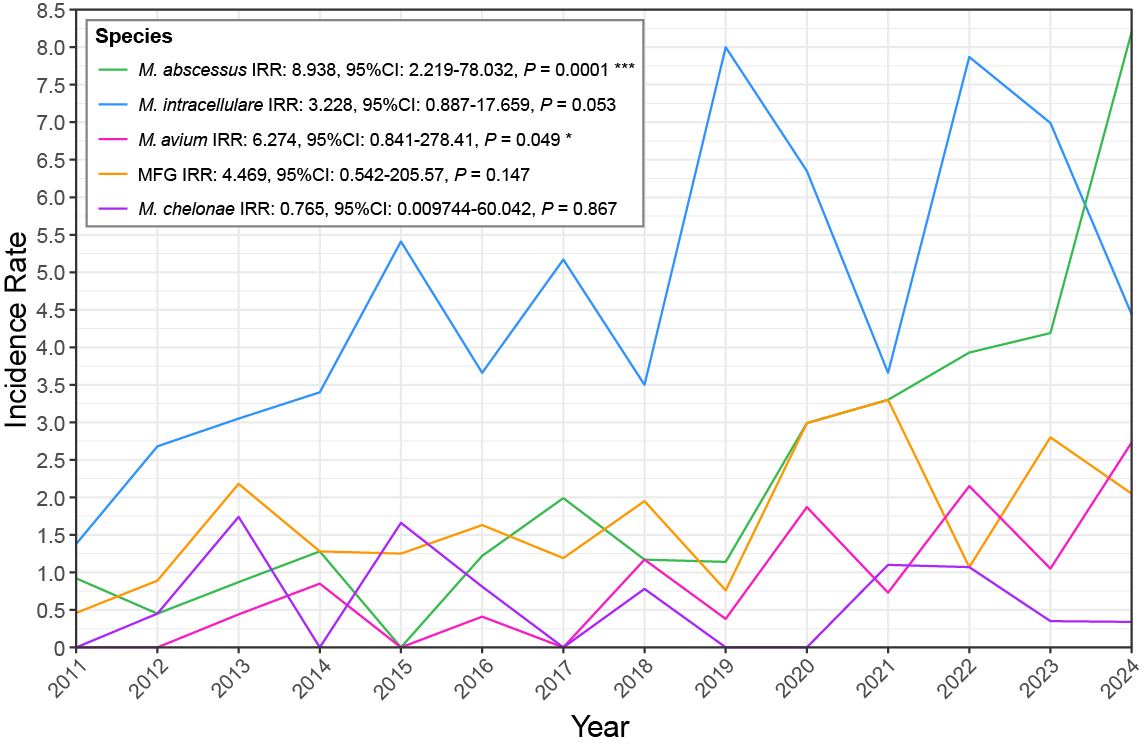

**Supplementary Figure 2. Incidence rate (per 100,000 First Nations people) of the top five nontuberculous mycobacterial species or species groups in First Nations people in Queensland.** MFG: *Mycobacterium fortuitum* Group; IRR: Incidence rate ratio; CI: Confidence interval. Significance is indicated by asterisks where *=*P*≤0.05 and ***=*P*≤0.001.
